# Predictive Autoantibodies Before the Diagnosis of Type I Diabetes in Adults

**DOI:** 10.64898/2026.06.23.26355661

**Authors:** Peter D. Burbelo, Robert Nee, Julio A. Huapaya, Richard Plasse, Myungjin Kim, Sarah Gordon, Giovanni Di Pasquale, John A. Chiorini, Stephen Olson

## Abstract

Recent epidemiologic studies indicate that adult-onset type 1 diabetes (AOT1D) is more common than childhood-onset type 1 diabetes, yet it remains clinically underrecognized. Because little is known about the emergence of islet autoantibodies in AOT1D, we conducted a retrospective study using electronic medical records from the United States Military Health System and longitudinal serum samples from 169 individuals with AOT1D and 40 healthy controls obtained from the Department of Defense Serum Repository. Among 643 prediagnostic samples from individuals with AOT1D, IA-2 autoantibodies were the most prevalent (50%), followed by GADA (46%), IA-2β (34%), ZnT8-R (27%), and ZnT8-W (15%). Overall, 85% (144/169) of subjects were seropositive for at least one autoantibody prior to diagnosis. Analysis of the earliest available sample from all of the AOT1D cases, grouped into 5-year intervals preceding diagnosis, demonstrated a progressive increase in seropositivity over time: 38% of subjects were seropositive more than 20 years before diagnosis, increasing to 44% at 20–15 years, 59% at 15–10 years, 73% at 10–5 years, and 91% within 5 years of diagnosis. Among the 144 seropositive individuals, positivity for two or more autoantibodies was the most common pattern, occurring in 50% (72/144) of cases. Isolated GADA positivity (22%) and isolated IA-2/IA-2β positivity (24%) occurred at similar frequencies, whereas isolated ZnT8 positivity was uncommon (4%). Temporal analysis showed that isolated GADA positivity appeared earliest, with a median onset of 7.9 years before diagnosis, whereas multiple-autoantibody positivity, IA-2 positivity, and ZnT8 positivity emerged later, with median onsets of 4.6, 4.5, and 1.9 years before diagnosis, respectively. These findings extend observations from pediatric type 1 diabetes to adults and demonstrate that AOT1D-associated autoimmunity often begins decades before clinical diagnosis, highlighting a potentially important window for risk stratification and preventive intervention.

## Introduction

Type 1 diabetes (T1D) is a chronic autoimmune disease caused by immune-mediated destruction of pancreatic β cells, with variability in genetic risk and age at onset [1]. While T1D is recognized as a T-cell-mediated disease, multiple contributing factors including genetic susceptibility, antiviral responses, and environmental factors may interact to drive β-cell destruction and influence disease onset and progression [2]. A key diagnostic and predictive tool for T1D is the presence of well-established islet autoantibodies including against islet cells (ICA), insulin (IAA), glutamic acid decarboxylase (GADA), insulinoma-associated protein 2 antigen (IA-2), and zinc transporter-8 (ZNT8) [3]. Importantly, there is substantial heterogeneity in the appearance of autoantibodies in T1D, and distinct disease endotypes have been proposed in part by their autoantibody profiles [4]. Autoantibodies can emerge in genetically susceptible infants as early as 6 months of age, with incidence peaking around 2 years of age [5, 6]. The early appearance of IAA is strongly associated with the HLA-DR4/DQ8 haplotype and tend to be diagnosed with T1D between 1 and 2 years of age. In contrast, children with GADA as the sole early autoantibody marker are more commonly associated with the HLA-DR3/DQ2 haplotype and tend to be diagnosed later, between 3 and 6 years of age [7, 8]. In children diagnosed with T1D at a later onset (after age 10), IAA is often absent or present at low titers [9], while GADA is often the most common persistent marker [10]. Lastly, the presence of multiple autoantibodies is the most definitive clinical marker of progression to T1D [11]. In addition to autoantibodies, further evidence for age-related differences comes from histopathologic studies identifying distinct endotypes based pancreatic immune infiltration [12, 13]. Children diagnosed before age 13 tend to exhibit CD20^high^ insulitis, characterized by CD8^+^ T-cell dominance and few residual β-cells. In contrast, children diagnosed at 13 or older showed CD20^low^ insulitis, with fewer CD8^+^ T-cells and more residual β-cells.

Despite these insights, our understanding of the natural history and endotypes of T1D have largely been shaped by studies focused on childhood-onset T1D [11, 14]. Recent epidemiologic studies have revealed that adult-onset type I diabetes (referred here as AOT1D) may be more prevalent than childhood-onset T1D and remains clinically underrecognized [15–18]. Emerging evidence suggests that AOT1D differs from childhood-onset T1D in its genetic, immunologic, and metabolic features, including a higher prevalence of protective genotypes [19, 20] and lower high-risk HLA heterozygosity [21]. A deeper understanding of the prodromal phase of autoimmunity, the emergence of islet autoantibodies, and the factors contributing to the delayed clinical onset of AOT1D compared to childhood-onset T1D could provide important insights into disease progression and therapeutic intervention strategies.

Here, we conducted a retrospective study using electronic medical records (EMRs) from the U.S. Military Health System (USMHS) and serial serum samples from the Department of Defense Serum Repository (DoDSR) to investigate the emergence and temporal trajectories of islet autoantibodies prior to the onset of AOT1D.

## Material and Methods

### Ethics statement

This retrospective study of AOT1D was approved by the Walter Reed National Military Medical Center Institutional Review Board (protocol 945439, “Prediagnostic Antibodies in Adult-Onset Autoimmune Diabetes and Long Term Outcomes”). The requirement for informed consent was waived by the Institutional Review Board because all data and samples were de-identified prior to analysis. No personally identifiable information was collected or disclosed. All procedures were conducted in accordance with the ethical standards of the National Institutes of Health and the Declaration of Helsinki.

### AOT1D cohort selection

The USMHS provides care for approximately 9.6 million beneficiaries and maintains a unified EMR system linked to a serum repository that has supported several prior epidemiologic serological studies [22–25].

In the retrospective study, we used EMRs from the USHMS together with longitudinal serum samples from the DoDSR to evaluate prediagnostic diabetes-associated autoantibodies in individuals who later developed AOT1D. Between 10/13/2019 to 2/10/2020, EMR data were accessed for research purposes to identify eligible AOT1D cases. The different serum samples used in this study were collected from active- and inactive-duty US military personnel between 1/01/1970, and 11/01/2019. As outlined in the attrition diagram (**Fig 1**), 3,332 potential cases were initially identified as having a diagnosis of AOT1D from EMRs between 2005 and 2019 using ICD-9 codes 250.x1 and 250.x3 and ICD-10 code E10. Eligibility and cohort inclusion were confirmed through manual EMR review and evaluation of available clinical autoantibody results, including ICA, IAA, GADA, IA-2, and ZnT8, resulting in 1,709 confirmed individuals with diabetes-associated autoantibodies. Using EMR data from the integrated USMHS from 2009 to 2018, we estimated an average annual AOT1D incidence of 9.8 cases per 100,000 (95% CI 6.2-13.2 per 100,000), based on 223 new cases per 2,285,914 individuals per year (**data not shown).** However, when limited to patients with confirmed islet autoantibodies seropositivity, the incidence dropped to 5.0 per 100,000 (95% CI 4.2-6.0 per 100,000) based on 115 new cases per 2,285,914 individuals per year (**data not shown**).

**Fig 1.**
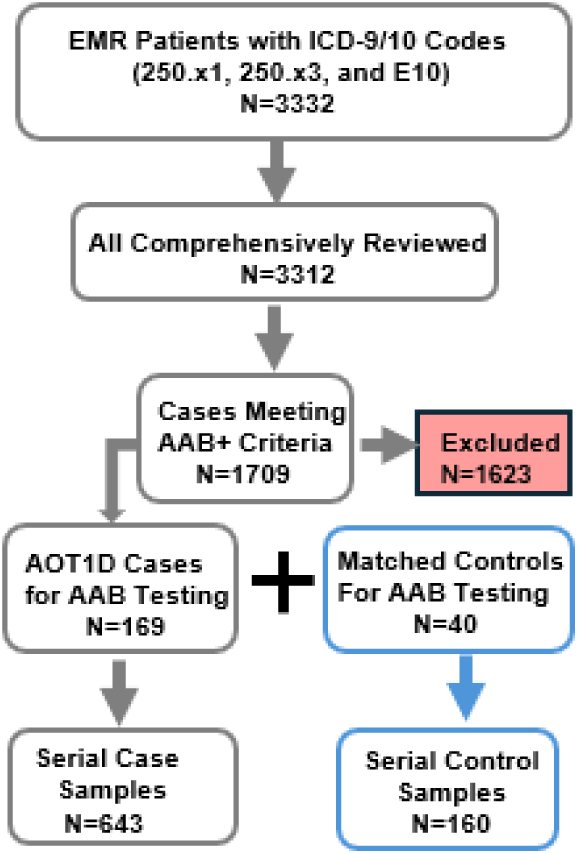
Selection of the AOT1D study cohort. Electronic medical records (EMRs) from the USHMS were used to identify 3,332 potential AOT1D cases diagnosed between 2005 and 2019 based on ICD-9 codes 250.x1 and 250.x3 and ICD-10 code E10. Manual review of EMRs and confirmation of islet autoantibody positivity resulted in 1,709 confirmed AOT1D cases. For the longitudinal analysis of prediagnostic islet autoantibodies, a subset of archived serum samples from 169 individuals with AOT1D and 40 matched healthy controls obtained from the DoDSR was included in the study.

Because the DoDSR limited the maximum number of 2,500 serum samples for our study, we initially retrieved a cohort collection that included 511 individuals with AOT1D and 126 matched healthy controls. Case selection was based on serum sample availability, the ability to determine the timing of insulin initiation, the number of islet autoantibodies present at diagnosis, availability of matched healthy controls, and the presence of ZnT8A testing data at diagnosis, which were limited in the EMR. Due to resource and personnel constraints, only a subset of banked serum samples from 169 individuals with AOT1D and 40 age and sex-matched healthy controls, obtained from the DoDSR, were used for autoantibody analysis (**Fig 1**). Clinical characteristics of the 169 AOT1D cases at diagnosis are shown in **Table 1**. Among the selected AOT1D cases, 140/169 (82.8%) had four longitudinal serum samples available, 27/169 (16.0%) had three samples, and 2/169 (1.2%) had only one sample. Most cases (124/169, 73.4%) had at least one serum sample collected within 1 year prior to diagnosis, and nearly all (166/169, 98.2%) had a sample within 2 years prior to diagnosis. Of the three cases without a sample within 2 years of diagnosis, two had only a single prediagnostic sample available. The cohort derived from the U.S. Military Health System was predominantly male (89.9%). The final testing cohort comprised 803 longitudinal serum samples, including 643 samples from AOT1D cases and 160 samples from matched healthy controls.

**Table 1.** Characteristics of the AOT1D Cohort.

|  |  |
| --- | --- |
|  | ALL (n=169) |
| Age (Median years) | 28 (24,35) |
| <35 y/o (%) | 73.4 (124/169) |
| Sex (% male) | 89.9 (151/169) |
| Race |  |
| Black (%) | 22.5 (38/169) |
| White (%) | 65.1 (110/169) |
| Other (%) | 12.4 (21/169) |
| GADA at Diagnosis (%) | 84.6 (143/169) |
| IA-2A at Diagnosis (%) | 75.2 (106/141) |
| ZnT8 at Diagnosis (%) | 73.9 (34/46) |

Available clinical data is shown for the AOT1D cohort. AOT1D cases were de-identified which prevented full data collection by chart review.

### Luciferase immunoprecipitation systems autoantibody testing

Luciferase immunoprecipitation systems (LIPS) assays using luciferase fusion proteins were employed to measure autoantibodies against GAD65, IA-2, IA-2β, ZnT8-R, and ZnT8-W, as previously described for islet autoantigens [26–28]. Insulin autoantibodies were not assessed because the assay was unavailable. For LIPS testing, 803 longitudinal serum samples were provided to a single investigator (PDB) in a blinded manner; participant identities, diagnoses, and the chronological order of serial samples were concealed throughout testing.

Autoantibody testing was performed using previously described methodology [29]. Briefly, plasmids encoding luciferase-antigen fusion proteins were transfected into Cos1 cells, and crude cell extracts were prepared 48 hours later. Serum samples were diluted 1:10 in assay buffer A (20 mM Tris, pH 7.5, 150 mM NaCl, 5 mM MgCl₂, 1% Triton X-100). Diluted serum (10 μl) was incubated in 96-well microtiter plates containing 40 μl of buffer A and 50 μl of Cos1 cell extract containing approximately 10^7^ light units (LU) of luciferase-antigen fusion protein extract.

Following incubation for 1 hour at room temperature, mixtures were transferred to filter plates containing protein A/G beads and incubated for an additional hour. Immune complexes were washed eight times with buffer A and twice with PBS to remove unbound antigen. Luciferase activity was measured following the addition of coelenterazine substrate (Promega, Fitchburg, WI) using a Berthold LB 960 Centro microplate luminometer (Berthold Technologies, Oak Ridge, TN). After completion of autoantibody testing, sample identities were unblinded.

Positivity thresholds for each autoantibody were established as the mean plus 3 standard deviations of LU values derived from the 160 healthy control samples. The complete dataset, including raw autoantibody values and antigen-specific cutoff-adjusted values for all AOT1D and control samples, is provided in the **Supplemental Data Set 1**.

## Data Analysis

GraphPad Prism software (GraphPad Software, San Diego, CA) was used for data visualization and statistical analyses. Comparisons of autoantibody levels between groups were performed using Mann–Whitney *U* tests. To facilitate comparison of autoantibody profiles in Figure 6, the measured values and positivity thresholds GADA, IA-2, and ZnT-8 autoantibodies were normalized to 10,000 light units (LU), in which only data for the ZnT8-R isoform is shown.

## Results

### Clinical characteristics of the AOT1D cohort

This retrospective study analyzed archived longitudinal serum samples from 169 subjects who subsequently developed AOT1D. The majority of subjects were White (65.1%) and male (89.9%) (**Table 1)**. As part of the inclusion criteria, all participants had evidence of islet-associated autoantibodies at diagnosis. The median age at diagnosis was 28 years, and 73.4% of subjects in the cohort developed AOT1D before the age of 35 years.

### Seropositive autoantibodies in AOT1D before diagnosis

Luciferase immunoprecipitation systems (LIPS) immunoassays were used to measure autoantibodies in blinded, longitudinal serum samples from the cohort (n = 803) against five diabetes-associated antigens: IA-2, IA-2β, GADA, and two ZnT8 variants, ZnT8-R and ZnT8-W. After un-blinding, cut-off values were assigned for each antigen and seropositivity status for each serum-antigen pair was established. Using these thresholds, assay specificity exceeded 98% for IA-2β, 99% for IA-2 and GADA, and 100% for ZnT8-R and ZnT8-W (**Fig 2 and Fig S1**).

**Fig 2.**
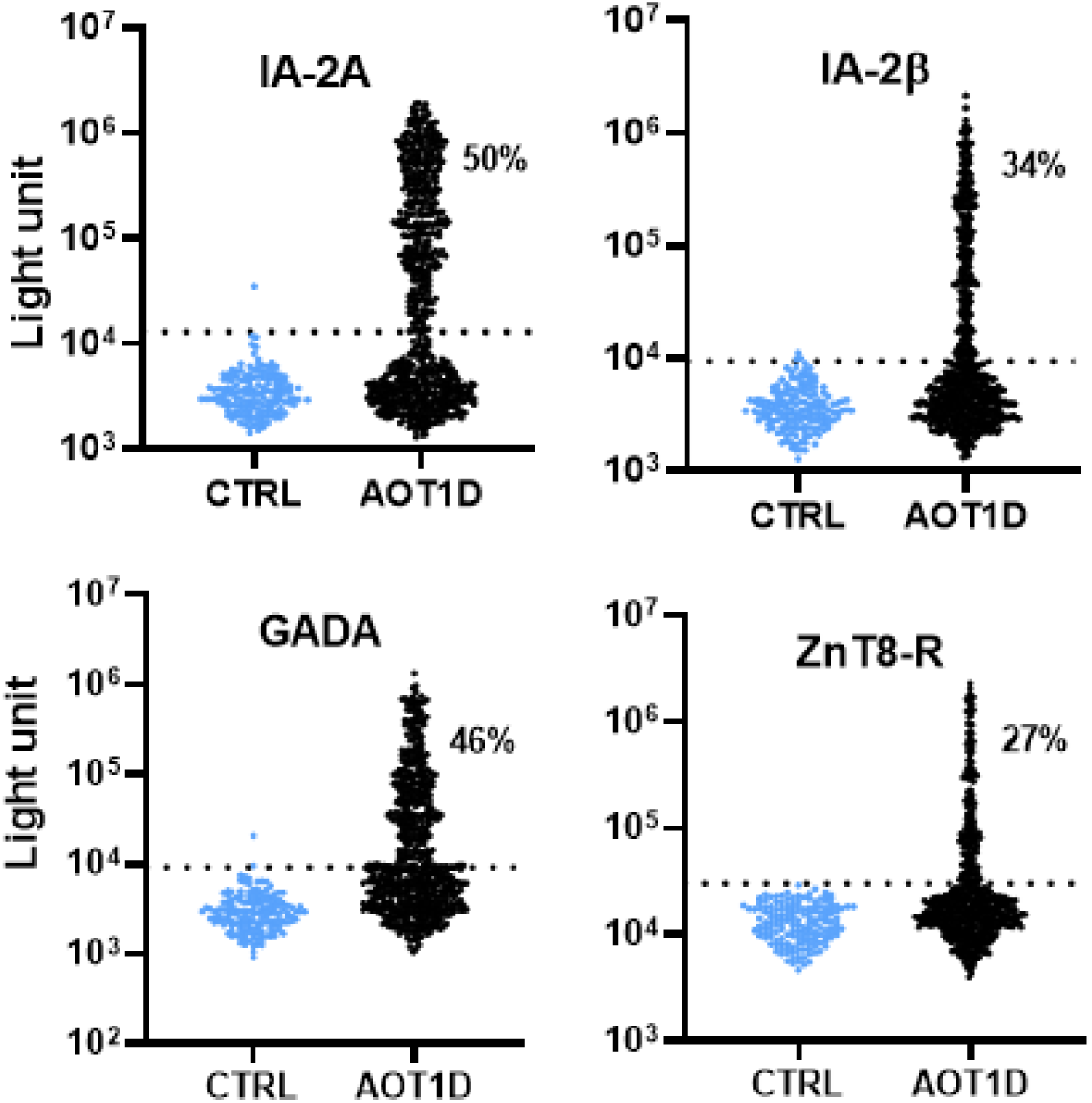
Detection and frequency of islet autoantibodies before diagnosis of AOT1D. Autoantibody levels were determined by LIPS against five islet autoantigens including (A) IA-2, (B) IA-2β, (C) GADA, and (D) ZnT-8R. Each symbol represents a serum sample from a healthy control subject (CTRL) or an individual who subsequently developed AOT1D. Autoantibody levels are expressed as light units (LU) and plotted on a log10 scale. Dashed lines indicate the cutoff values for seropositivity, defined as the mean plus 3 standard deviations of the healthy controls. Percentages indicate the frequency of seropositive samples for each autoantigen.

Among the 643 prediagnostic AOT1D samples, IA-2 autoantibodies were the most prevalent (50%), followed by GADA (46%), IA-2β (34%), ZnT8-R (27%), and ZnT8-W (15%). Overall, 85% (144/169) of the AOT1D cases were seropositive for at least one autoantibody before diagnosis.

### A majority of AOT1D cases show seropositivity a decade or more before diagnosis

A majority of AOT1D cases exhibited seropositivity a decade or more before diagnosis. Because the first available longitudinal serum sample for each subject in the AOT1D cohort (n = 169) was collected at varying times before diagnosis, samples were grouped into 5-year intervals for analysis. This analysis showed that the proportion of subjects positive for at least one autoantibody increased progressively as the diagnosis date approached (**Fig. 3**). Specifically, 38% of subjects were seropositive more than 20 years before diagnosis, 44% at 15–20 years, 59% at 10–15 years, 73% at 5–10 years, and 91% within 5 years of diagnosis.

**Fig 3.**
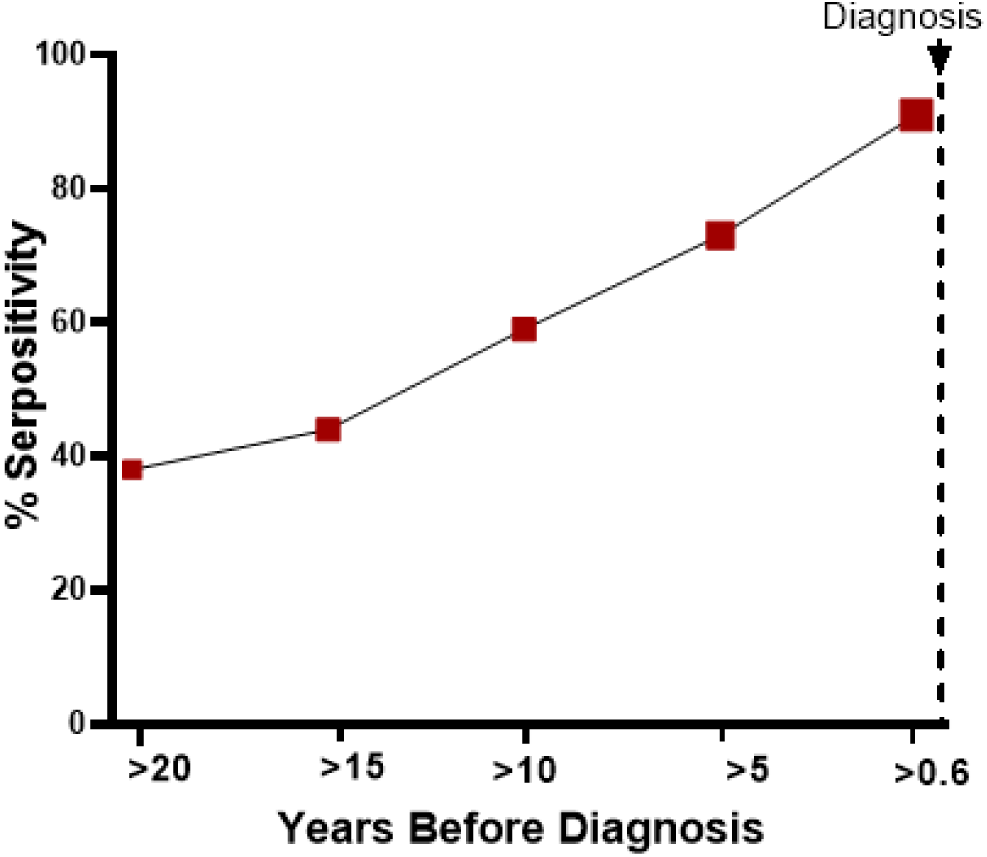
Frequency of islet autoantibody positivity before AOT1D diagnosis. Because the timing of the earliest available longitudinal serum sample varied among subjects in the AOT1D cohort (n = 169), samples were grouped into 5-year intervals relative to diagnosis (x-axis). The y-axis shows the proportion of subjects positive for at least one islet autoantibody within each interval. Red squares indicate the number of subjects analyzed in each interval: >20 years (n = 13), 15–20 years (n = 25), 10–15 years (n = 32), 5–10 years (n = 45), and <5 years before diagnosis (n = 54).

### Seropositivity at the earliest available sample

Closer examination of the 144 seropositive AOT1D cases revealed that 19% (28/144) were seronegative at their earliest available longitudinal sample; these samples were collected at a median of 14.7 years before diagnosis (**Fig. 4**). In contrast, the remaining 81% of cases were already seropositive at their earliest available sample, which was collected at a median of 6.2 years before diagnosis. Notably, some individuals in this group exhibited seropositivity more than 20 years before diagnosis, with the earliest detected case occurring 27 years prior to diagnosis. Because most individuals in this group were already seropositive for at least one autoantibody in their earliest available sample, the observed intervals likely underestimate the true duration between initial seroconversion and clinical diagnosis.

**Fig 4.**
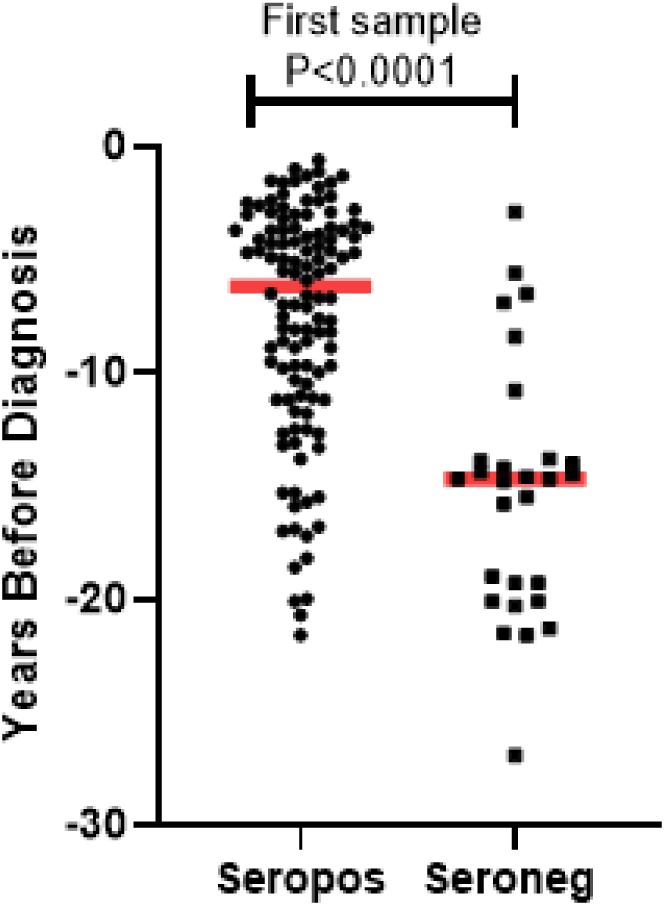
Islet autoantibody status at the first available blood draw before AOT1D diagnosis. Subjects were classified as seropositive (Seropos) or seronegative (Seroneg) based on reactivity to any of the five islet autoantibodies measured in the earliest available serum sample. The timing of the first available sample relative to diagnosis is shown for each subject. Red bars indicate the median time before diagnosis for each group. Statistical significance was assessed using the Mann–Whitney *U* test.

### Autoantibody Patterns at the First Appearance of Seropositivity

A detailed analysis of autoantibody patterns at the first appearance of seropositivity among the 144 seropositive AOT1D cases revealed that positivity for two or more islet autoantibodies was the most common pattern, occurring in 50% (72/144) of cases (**Fig. 5**). Isolated GADA seropositivity (22%, 32/144) and isolated IA-2/IA-2β seropositivity (24%, 34/144) occurred at similar frequencies at the first seropositive time point, whereas isolated ZnT8 seropositivity was uncommon (4%).

**Fig 5.**
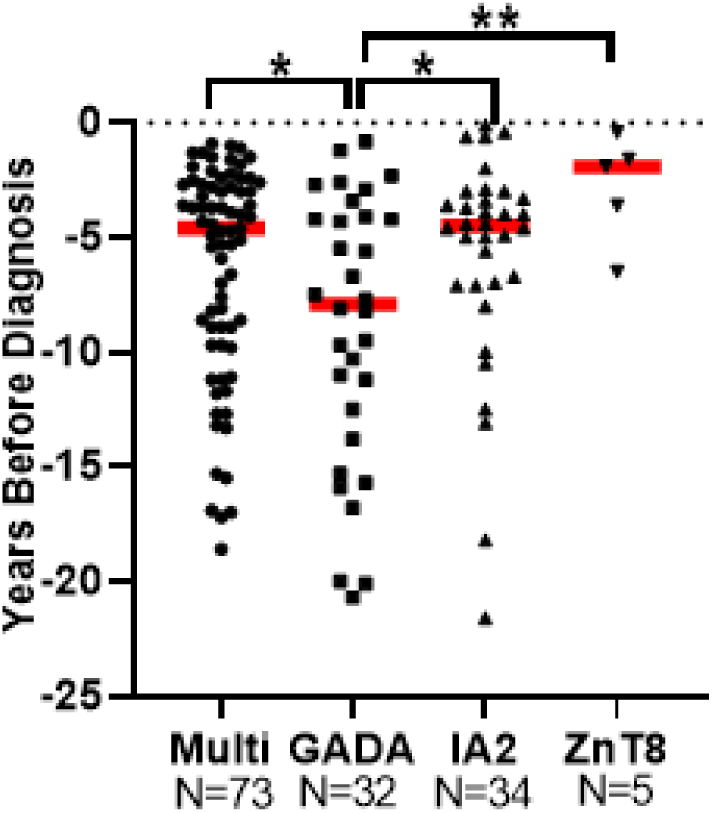
Autoantibody patterns at the earliest seropositive sample before AOT1D diagnosis. The autoantibody pattern detected in the earliest seropositive serum sample from each subject is plotted according to the number of years before diagnosis that the sample was collected. Subjects seropositive for two or more autoantibodies were grouped in the Multi category. For this analysis, IA-2A and IA-2β reactivity were combined and were not considered separate autoantibodies when determining multiple-autoantibody positivity. Red bars indicate the median time before diagnosis for each autoantibody pattern. Only statistically significant comparisons are shown and were assessed using the Mann–Whitney U test (p < 0.05, **p ≤ 0.01).

**Fig 6.**
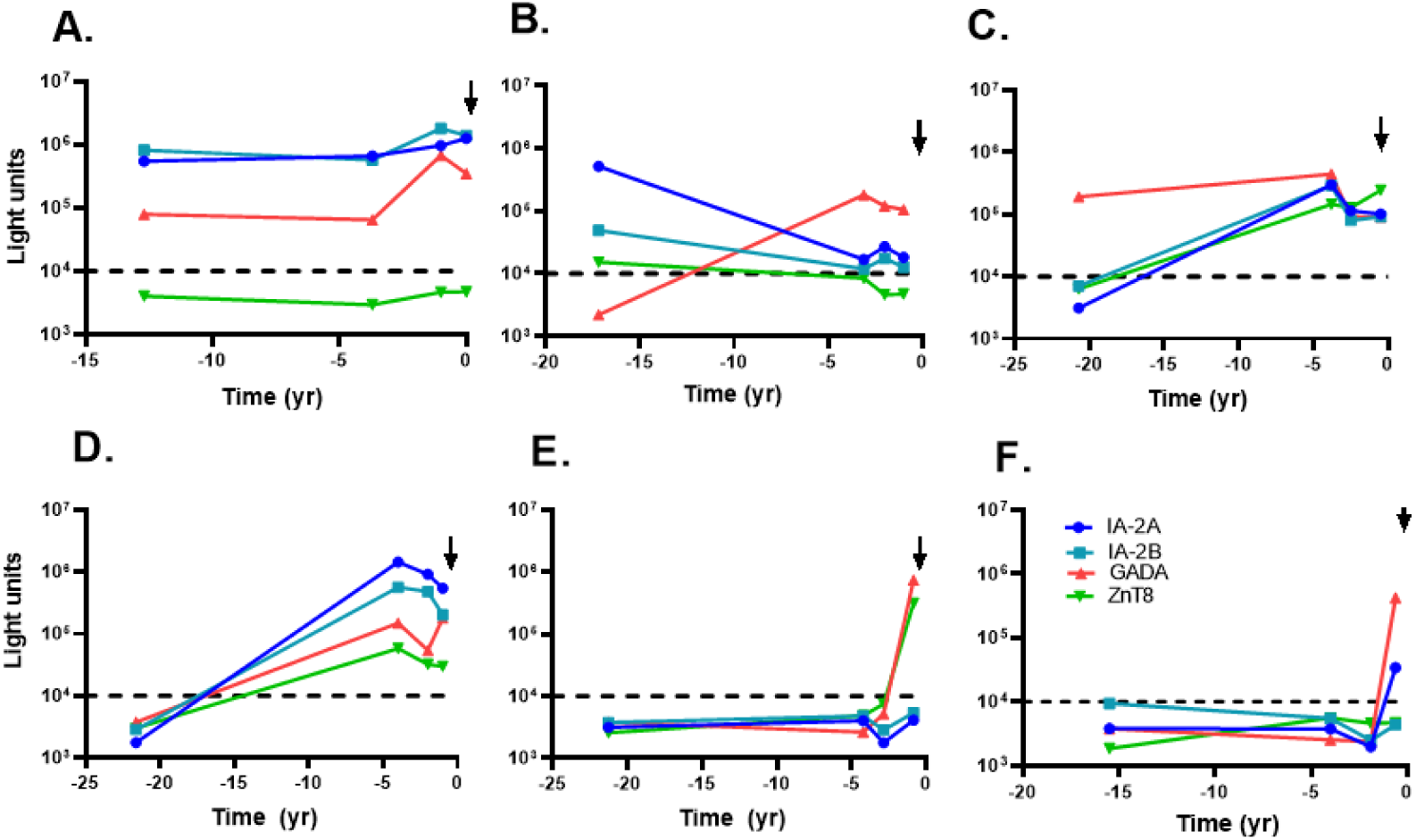
Representative longitudinal prediagnostic islet autoantibody profiles in AOT1D. Representative examples of longitudinal autoantibody profiles from AOT1D subjects who were seropositive (A–C) or seronegative (D–F) at their earliest available serum sample. Time zero, indicated by the vertical black arrow, denotes the time of AOT1D diagnosis. Autoantibody levels for IA-2, IA-2β, GADA, and ZnT8-R are expressed as light units (LU) and plotted on a log10 scale (y-axis). Dotted black lines indicate the common cutoff value used to define seropositivity for the different autoantibodies. Notably, two subjects (E and F) remained seronegative until their fourth serial sample and became seropositive within the final year before AOT1D diagnosis.

Further analysis of autoantibody patterns at the first seropositive sample revealed significant differences in their timing of appearance. GADA appeared earliest, with a median detection time of 7.9 years before diagnosis. In contrast, multiple-autoantibody positivity, IA-2/IA-2β autoantibodies, and ZnT8 autoantibodies emerged later, at median times of 4.6, 4.5, and 1.9 years before diagnosis, respectively (Fig. 5). The differences in timing between GADA and multiple-autoantibody positivity (Mann–Whitney U test, *P* = 0.04), GADA and IA-2/IA-2β positivity (*P* = 0.03), and GADA and ZnT8 positivity (*P* = 0.01) were statistically significant.

Although the timing of AOT1D serum samples provided by the DODSR varied among subjects, several distinct prediagnostic autoantibody patterns emerged (**Fig. 6**). One common pattern was persistent seropositivity, often lasting more than a decade and frequently characterized by reactivity to multiple islet autoantibodies (**Fig. 6A, B**). Another pattern involved isolated early GADA seropositivity that either remained restricted to GADA or was followed by the appearance of additional autoantibodies over time (**Fig. 6C**). Among the minority of cases that were seronegative at their earliest available samples (**Fig. 6D-F**), some later developed multiple islet autoantibodies across successive time points (**Fig. 6D**). In contrast, two subjects remained seronegative until their final available sample, collected approximately 6 months before diagnosis, when they became seropositive (**Fig. 6E, 6F**). These cases suggest the possibility of a more rapid disease onset than the more prolonged trajectories observed in most subjects.

## Conclusion

In this study, we characterized the longitudinal development of islet autoantibodies in individuals who subsequently developed AOT1D using serial serum samples collected years before diagnosis. Prediagnostic islet autoantibodies were detected in 86% of cases, demonstrating that humoral autoimmunity is present in most individuals before the clinical onset of disease. In many subjects, autoantibody responses were detected repeatedly across multiple longitudinal samples, indicating sustained humoral autoimmunity before clinical onset. Moreover, the prevalence of autoantibody positivity increased steadily as diagnosis approached, and 38% of subjects were seropositive more than 20 years before diagnosis. These findings indicate that AOT1D is preceded by a prolonged period of progressive autoimmune activity and extend observations from pediatric cohorts showing that islet autoantibodies may precede type 1 diabetes by many years [11]. Our results provide direct evidence that an even longer extended preclinical phase is a characteristic feature of adult-onset autoimmune disease.

Although insulin autoantibodies were not assessed, distinct temporal patterns were observed among the remaining major islet autoantibodies. GADA was typically the earliest detected autoantibody, appearing a median of approximately 8 years before diagnosis, whereas IA-2A and ZnT8 autoantibodies generally emerged closer to disease onset. In addition, individuals who first exhibited isolated GADA positivity appeared to progress more slowly than those with IA-2A or multiple-autoantibody positivity. These findings are consistent with studies in children showing that IA-2A positivity and multiple-autoantibody positivity are associated with more rapid progression to type 1 diabetes [30]. We also observed substantial heterogeneity in disease trajectories, with some individuals remaining seropositive for decades before diagnosis and others developing autoantibodies only shortly before disease onset. Together, these observations suggest that different autoantibody profiles may reflect distinct stages and rates of disease progression in adults.

A major strength of this study is the use of longitudinal serum samples collected over many years before diagnosis, providing a unique view of the natural history of islet autoimmunity preceding AOT1D. Previous studies of AOT1D have largely relied on cross-sectional assessment of autoantibodies at or near diagnosis [31, 32]. However, several limitations should be considered. Information regarding family history, genetic susceptibility, and environmental exposures was incomplete, limiting our ability to identify factors influencing disease progression. In addition, sampling intervals varied among individuals, and the number of archived samples per subject was limited, preventing precise determination of seroconversion in some cases. Finally, insulin autoantibodies were not evaluated and therefore could not be incorporated into analyses of autoantibody trajectories.

Despite these limitations, our findings have important implications for the early identification of adults at risk for type 1 diabetes. The prolonged preclinical phase observed in many individuals suggests that targeted screening strategies in adults merit further evaluation, particularly approaches focused on GADA and IA-2, which were among the earliest and most prevalent autoantibodies detected before diagnosis. Support for this strategy comes from a Finnish study in high-risk individuals, in which screening for GADA and IA-2 identified a substantial proportion of those who later developed type 1 diabetes over long-term follow-up [33]. Future studies integrating autoantibody measurements with genetic, proteomic, metabolomic, and other biomarkers may improve prediction of disease progression. As disease-modifying therapies such as teplizumab demonstrate the ability to delay progression to clinical type 1 diabetes [34], identifying adults during the earliest stages of autoimmune disease may become increasingly important. Expanding screening and prevention efforts to include adults could help preserve residual β-cell function and ultimately delay or prevent the onset of insulin-dependent diabetes.

## Disclaimer

The contributions of the NIH authors are considered Works of the United States Government. The findings and conclusions presented in this paper are those of the authors and do not necessarily reflect the views, official policy or positions of the NIH, the U.S. Department of Health and Human Services, the Defense Health Agency, Department of War, nor the U.S. Government. The identification of specific products or scientific instrumentation is considered integral to the scientific endeavor and does not constitute endorsement or implied endorsement on part of the authors, Department of War or any component agency.

## Data Availability

All data produced in the present work are contained in the manuscript.

